# Workplace Infection Prevention Control Measures and Work Engagement During the COVID-19 Pandemic among Japanese Workers: A Prospective Cohort Study

**DOI:** 10.1101/2022.04.11.22273753

**Authors:** Kazunori Ikegami, Hajime Ando, Yoshihisa Fujino, Hisashi Eguchi, Keiji Muramatsu, Tomohisa Nagata, Seiichiro Tateishi, Mayumi Tsuji, Akira Ogami, the CORoNaWork project

## Abstract

**Objectives:** It has been widely reported that the COVID-19 pandemic may have a psychological influence on people. Thus, it could be important to note how workplace infection prevention and control (IPC) measures for COVID-19 contribute to positive mental health among workers. We hypothesized that if workplace IPC measures are adequately implemented, they would have a positive effect on employees’ work engagement.

**Methods:** We conducted an internet-based prospective cohort study from December 2020 (baseline) to December 2021 (follow-up after one year) using self-administered questionnaires. At baseline, 27,036 workers completed the questionnaires, while 18,560 (68.7%) participated in the one-year follow-up. After excluding the 6,578 participants who changed jobs or retired during the survey period, or telecommuted more than four days per week, 11,982 participants were analyzed. We asked participants about the implementation of workplace IPC measures at baseline and conducted a nine-item version of the Utrecht Work Engagement Scale (UWES-9) at follow-up.

**Results:** Four groups were created according to the number of workplace IPC measures implemented. The mean (SD) UWES-9 score of the “0–2” group was the lowest at 18.3 (13.2), while that of the “8” group was the highest at 22.6 (12.6). The scores of the “3– 5,” “6–7,” and “8” groups were significantly higher than that of the “0–2” group (all, p<0.001). The p trend of the four groups was also significant (p<0.001).

**Conclusions:** Promoting workplace IPC measures improves workers’ work engagement, and a dose-response relationship exists between workplace IPC measures and work engagement.

## Introduction

The coronavirus disease (COVID-19), which was caused by severe acute respiratory syndrome coronavirus 2 (SARS-CoV-2) and broke out in December 2019, has caused a pandemic around the world due to its viral mutations, and has yet to be fully contained.^1, 2^ In Japan, a COVID-19 pandemic occurred after 2020, and the Japanese government repeatedly declared a state or quasi-state of emergency; focused on anti-infection measures; and strengthened infection control measures such as non-pharmaceutical interventions (NPIs). In addition, COVID-19 vaccination was also promoted both in the community and in occupational fields. However, in November 2021, the SARS-CoV-2 omicron variant (B.1.1.529) was classified as a variant of concern (VOC); by March 2022, the omicron variant continued to spread COVID-19 worldwide, including in Japan. Hence, the infection prevention control (IPC) of COVID-19 has been an important issue.^3^

COVID-19 is thought to be transmitted mainly by droplets containing the virus (droplet infection).^4^ However, it has been reported that droplet nuclei (aerosols), which are transformed when droplets float in the air, can also cause COVID-19 infection.^5, 6^ Various NPI strategies have been implemented to reduce SARS-CoV-2 transmission, including mask-wearing, hand hygiene, physical distancing, and proper room ventilation.^7^ In particular, the workplace is considered one of the most likely places for the spread of COVID-19 because many employees work and communicate in the same space.

In Japan, workplace IPC measures are one of the most important issues in fulfilling a company’s obligation to promote employees’ health and safety and ensure business continuity amidst the COVID-19 epidemic. To this end, many guidelines and checklists for workplace IPC measures have been published, including “A Guide for Businesses and Employers Responding to Novel Coronavirus Disease (COVID-19),” which was published by the Japanese Society for Occupational Health (JSOH).^8^ Based on these guidelines, workplace IPC measures have been implemented in many companies. Indeed, in addition to basic measures such as physical distancing, wearing masks, and washing hands, other proposed measures include enhancing office room ventilation; refraining from or restricting business trips, visitors, social gatherings, and face-to-face meetings; setting up partitions; daily physical condition checks; and promoting sick leave when employees feel ill.^8^

It has been widely reported that the COVID-19 pandemic may have an important psychological influence on people.^9^ We are particularly interested in the impact of workplace IPC measures on workers’ mental health. In general, work environment, work organization, and work-related behaviors are considered to be factors that influence workers’ mental health, psychological distress, and well-being.^10^ There are also reports on the COVID-19 pandemic and work stress. For example, it has been reported that anxiety about COVID-19 infection in the workplace may enhance job demands and psychological distress. ^11^ It has likewise been reported that telecommuting, which is implemented as a COVID-19 IPC measure, has a positive impact on workers’ work engagement.^12^ Thus, it is important to clarify how workplace environmental factors and work-related behaviors affect workers’ mental health during the COVID-19 pandemic.

In recent years, mental health support for workers has come to be regarded as important—not only in preventing workers’ mental disorders and resolving their mental problems, but also in promoting the revitalization of both workers and company organizations.^13^ Recent research in the field of occupational health has focused on themes involving positive mental health, such as improving well-being and productivity, as well as themes involving negative mental health, such as reducing job stress and treating depression.^14^ One of the leading indicators of positive mental health among workers that has been attracting attention is work engagement. Work engagement has been defined as “a positive, fulfilling, work-related state of mind that is characterized by vigor, dedication, and absorption.”^15, 16^ Employees with high work engagement are considered to be physically and mentally healthy, energetic, enthusiastic, and productive.^15, 17^ Work engagement can be easily assessed using questionnaires, and one such well-known questionnaire is the Utrecht Work Engagement Scale (UWES) developed by Schaufeli et al. The UWES has been standardized in many countries and has been confirmed to have good results in terms of reliability and validity.^15^ A Japanese version of UWES, which was developed by Shimazu et al.,^18^ has been used in many studies.

We hypothesized that if workplace IPC measures are adequately implemented, they would have a positive effect on employees’ work engagement. Therefore, in this study, we prospectively evaluated the influence of workplace IPC measures on workers’ work engagement by analyzing data from the Collaborative Online Research on the Novel-coronavirus and Work (CORoNaWork) Project.

## Methods

### Study Design and Setting

This study is a prospective cohort study conducted from December 2020 (baseline survey) to December 2021 (follow-up survey). Both the baseline and follow-up surveys were conducted using self-administered questionnaires on the Internet. All participants gave informed consent to participate in the study. The study was approved by the ethics committee of the University of Occupational and Environmental Health, Japan (Reference No. R2-079 and R3-006). The study protocol of the CORoNaWork study, including the sampling plan and subject recruitment procedure according to the Checklist for Reporting Results of Internet E-Surveys (CHERRIES) Checklist, has been reported in our previous work.^19-21^

The baseline survey was conducted when Japan was on maximum alert levels at the beginning of the third wave of COVID-19, as the number of COVID-19 infections and deaths were overwhelmingly higher in the third wave than in the first and second. The follow-up survey was conducted when the fifth wave had settled down and the number of infections was decreasing.

### Participants

#### Baseline survey

The target population was workers between the ages of 20 and 65 who were working at the time of the baseline survey. Sampling was conducted with allocation by region, occupation, and sex. Regions were divided into five levels of 47 prefectures according to the level of infection. Occupations were likewise divided into office workers and non-office workers. Thus, a total of 20 blocks of 5 regions, 2 occupations, and 2 sexes were assigned, and each block was sampled in equal numbers. We planned to study 30,000 people overall, and thus attempted to gain at least 1,500 participants in each block.

The survey was commissioned by Cross Marketing Inc. (Tokyo, Japan). Of their 4.7 million pre-registered monitors, approximately 600,000 were sent an email request to participate in the survey. Of these, 55,045 participated in the initial screening survey, while 33,087 met the inclusion criteria for the same.

Of the 33,087 initial participants, 27,036 (excluding those judged as fraudulent responses) were included in this analysis. The following criteria (i.e., the exclusion criteria) were used to determine fraudulent responses: extremely short response time (≤6 minutes), extremely low body weight (<30 kg), extremely short height (<140 cm), inconsistent answers to similar questions throughout the survey (e.g., inconsistency to questions about marital status and living situation), and wrong answers to a staged question used to identify fraudulent responses (choose the third-largest number from five numbers) (Figure 1).

**Figure 1.**
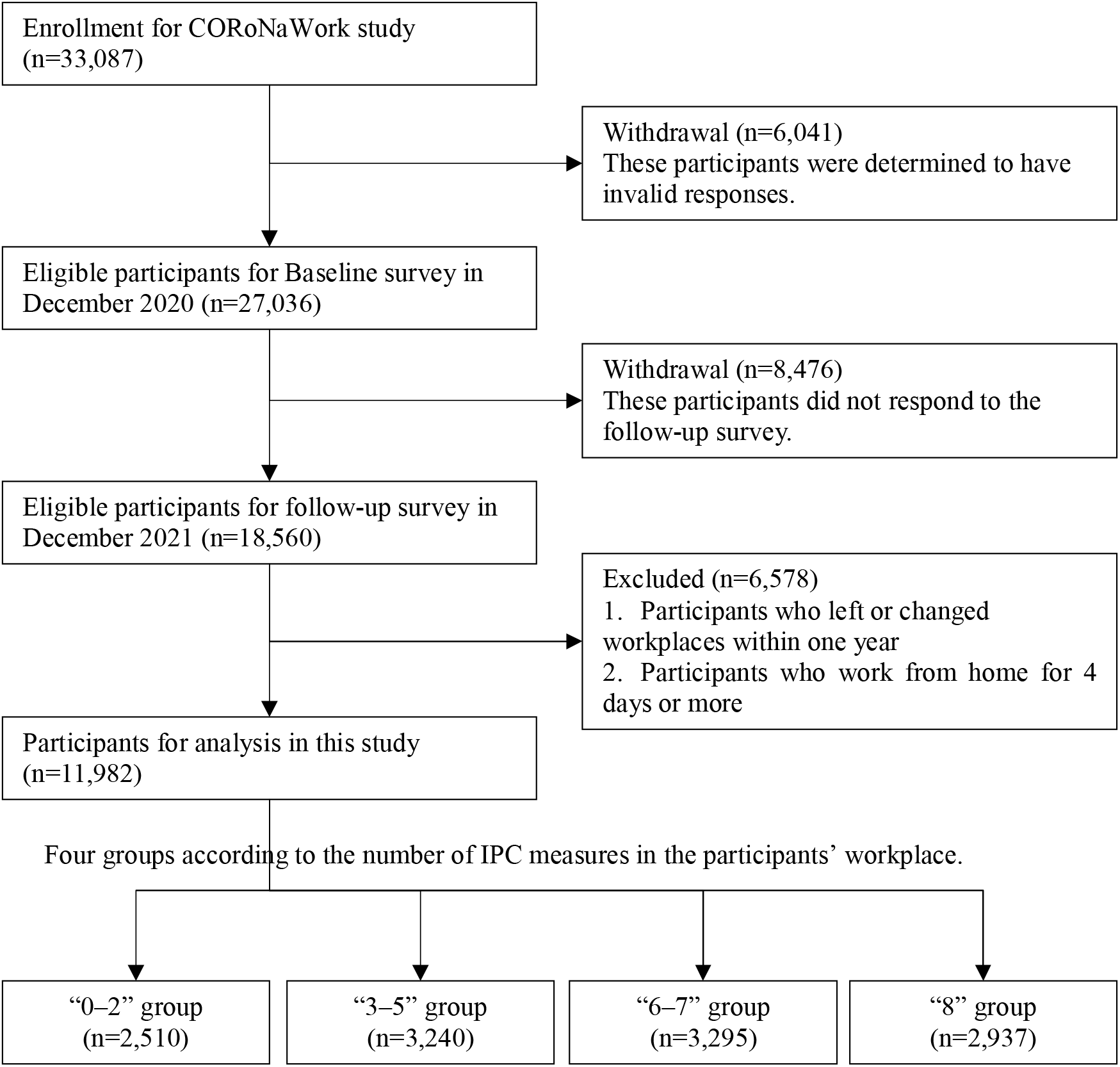
Flow chart of the study population selection.

#### Follow-up survey

A follow-up survey was conducted in December 2021, one year after baseline. A total of 18,560 (tracking rate: 68.7%) participated in the follow-up survey. We excluded 6,578 participants who changed jobs or retired during the survey period and those who telecommuted for more than four days per week (i.e., those who rarely worked in the workplace). Ultimately, 11,982 participants were analyzed (Figure 1).

### Evaluation of Work Engagement

A nine-item version of the UWES was used to measure work engagement.^22^ The Japanese version of the UWES-9 has been verified for reliability and validity by Shimazu et al.^23^ Each question item consists of a seven-point Likert scale ranging from 0 for “never” to 6 for “always.” The UWES-9 calculates three subscales (vitality, enthusiasm, and immersion), consisting of three items each, in addition to the total score. Higher scores indicate a higher state of work engagement. The score range of the UWES-9 is 0–54, and the range of each subscale is 0-18. In the present sample, the Cronbach’s alpha of UWES-9 (total score), vigor, dedication, and absorption were 0.97, 0.93, 0.91, and 0.92 respectively.

### Evaluation of Workplace IPC Measures for COVID-19

We asked the participants to answer whether or not the following eight workplace IPC measures for COVID-19 had been implemented by their workplace: 1. placing restrictions of business trips or going out for business (stopping business trips); 2. refraining from and placing restrictions on visitors (arranging health screenings for visitors); 3. refraining from or requesting a limit on the number of people at social gatherings and dinners (restricting work_related social gatherings and entertainment); 4. refraining from or limiting face-to-face internal meetings (restricting face-to-face meetings); 5. wearing masks at all times during work hours (encouraging mask-wearing at work); 6. installing partitions and revising the workplace layout (installing partitions or changing the working environment); 7. recommending workers perform daily temperature checks at home (enforcing temperature measurement); and 8. requesting employees not to come to work when they are not feeling well (requesting that employees refrain from going to work when ill). Variables regarding the eight items were calculated by totaling the number of “yes” responses for each participant (range: 0–8).

### Outcome and Measurements

The participants’ UWES-9 scores in the follow-up survey were used as outcome variables. The participants were divided into four groups (“0–2,” “3–5,” “6–7,” and “8”) according to the number of workplace IPC measures implemented in their workplace, and these were used as exposure variables.

Sex, age (20–29 years, 30–39 years, 40–49 years, 50–59 years, ≥60 years), educational background (middle school/high school, junior college/vocational school, university/graduate school), number of household members (1 person, 2 people, 3 people, ≥4 people), standard industrial classification (primary industry, secondary industry, tertiary industry), job type (regular employee, managers, others), and size of the workplace (1–9 employees, 10–49 employees, 50–999 employees, ≥1000 employees) were used as confounders. The standard industrial classification was defined by the Japanese Ministry of Internal Affairs and Communications. These variables, except for the size of the workplace, were collected in the baseline survey. While data on the size of the participants’ companies were collected in the baseline survey, the size of the workplace was likewise asked in the follow-up survey to obtain more detailed information.

### Statistical Analyses

To estimate whether the workplace IPC measures were associated with work engagement among the participants, we used a multilevel regression analysis nested in the prefecture of residence in order to account for regional variability. An age-sex adjusted model and multivariate-adjusted model were estimated. Both models included age, sex, education, number of household members, and the four groups according to the number of COVID-19 infection control measures in the workplace as the fixed effects, while the prefecture of residence was the random effect. In addition, the p-values of multilevel regression analysis were calculated by considering each category scale of the number of workplace IPC measures as continuous variables (p for trend). In all tests, the threshold for significance was set at p<0.05. Stata/SE Ver.15.1 (StataCorp LLC, College Station, Texas, United States) was used for the analysis.

## Results

### Participants and Descriptive Data

Compared to the participants working in workplaces with fewer workplace IPC measures, the participants working in companies with more workplace IPC measures tended to have higher educated and be married. Smaller-sized workplaces tended to have fewer workplace IPC measures, while larger-sized workplaces tended to have more. In terms of standard industry classification, the primary industry had the highest proportion of workplaces implementing “0–2” workplace IPC measures among the three industries, and the secondary industry had the highest proportion of workplaces implementing 8 workplace IPC measures. The tertiary industry had the highest proportion of workplaces implementing “3–5” and “6–7” workplace IPC measures (Table 1).

**Table 1.** Characteristics of each group according to the number of workplace IPC measures for COVID-19 at baseline. IPC, infection prevention and control.

| Item | Group by # of workplace IPC measures |  |  |  |  |  |  |  |
| --- | --- | --- | --- | --- | --- | --- | --- | --- |
|  | “0–2” group<br>(n=2,510) |  | “3–5” group<br>(n=3,240) |  | “6–7” group<br>(n=3,295) |  | “8” group<br>(n=2,937) |  |
|  | n | (%) | n | (%) | n | (%) | n | (%) |
| Sex, male | 1,507 | (60.0) | 1,764 | (54.4) | 1,843 | (55.9) | 1,686 | (57.4) |
| Age |  |  |  |  |  |  |  |  |
| 20–29 years | 93 | (3.7) | 168 | (5.2) | 159 | (4.8) | 159 | (5.4) |
| 30–39 years | 374 | (14.9) | 511 | (15.8) | 522 | (15.8) | 473 | (16.1) |
| 40–49 years | 839 | (33.4) | 1,068 | (33.0) | 1,039 | (31.5) | 886 | (30.2) |
| 50–59 years | 899 | (35.8) | 1,163 | (35.9) | 1,261 | (38.3) | 1,122 | (38.2) |
| ≥60 years | 305 | (12.2) | 330 | (10.2) | 314 | (9.5) | 297 | (10.1) |
| Education |  |  |  |  |  |  |  |  |
| Junior high / high school | 970 | (38.6) | 923 | (28.5) | 825 | (25.0) | 614 | (20.9) |
| Vocational school / college | 609 | (24.3) | 763 | (23.5) | 686 | (20.8) | 624 | (21.2) |
| University / graduate school | 931 | (37.1) | 1,554 | (48.0) | 1,784 | (54.1) | 1,699 | (57.8) |
| Size of workplace* |  |  |  |  |  |  |  |  |
| ≤9 employees | 1,392 | (55.5) | 995 | (30.7) | 578 | (17.5) | 330 | (11.2) |
| 10–49 employees | 664 | (26.5) | 1,121 | (34.6) | 1,027 | (31.2) | 682 | (23.2) |
| 50–999 employees | 381 | (15.2) | 943 | (29.1) | 1,357 | (41.2) | 1,416 | (48.2) |
| ≥1000 employees | 73 | (2.9) | 181 | (5.6) | 333 | (10.1) | 509 | (17.3) |
| Standard industrial classification† |  |  |  |  |  |  |  |  |
| Primary industry | 65 | (2.6) | 26 | (0.8) | 13 | (0.4) | 7 | (0.2) |
| Secondary industry | 759 | (30.2) | 693 | (21.4) | 779 | (23.6) | 919 | (31.3) |
| Tertiary industry | 1,686 | (67.2) | 2,521 | (77.8) | 2,503 | (76.0) | 2,011 | (68.5) |
| Job type |  |  |  |  |  |  |  |  |
| Regular employee | 1,289 | (51.4) | 1,488 | (45.9) | 1,485 | (45.1) | 1,383 | (47.1) |
| Manager | 138 | (5.5) | 280 | (8.6) | 430 | (13.1) | 500 | (17.0) |
| Other | 1,083 | (43.1) | 1,472 | (45.4) | 1,380 | (41.9) | 1,054 | (35.9) |
| Marital status, unmarried | 1,203 | (47.9) | 1,462 | (45.1) | 1,336 | (40.5) | 1,087 | (37.0) |
| # of household members |  |  |  |  |  |  |  |  |
| 1 person | 523 | (20.8) | 644 | (19.9) | 638 | (19.4) | 520 | (17.7) |
| 2 people | 710 | (28.3) | 883 | (27.3) | 883 | (26.8) | 728 | (24.8) |
| 3 people | 637 | (25.4) | 844 | (26.0) | 827 | (25.1) | 758 | (25.8) |
| ≥4 people | 640 | (25.5) | 869 | (26.8) | 947 | (28.7) | 931 | (31.7) |

IPC, infection prevention and control.
\* This standard industrial classification was defined by the Japanese Ministry of Internal Affairs and Communications.
† Although the data on the size of the companies where the participants worked was collected in the baseline survey, the size of the workplace was also collected in the follow-up survey to obtain more detailed information.

### UWES-9 Among Four Groups According to the Number of Workplace IPC Measures

As for the mean (SD) UWES-9 scores among the four groups according to the number of workplace IPC measures, the “0–2” group had the lowest at 18.3 (13.2), and the “8” group had the highest at 22.6 (12.6). In both the sex-age adjusted model and the multivariate model, the scores of the “3–5,” “6–7,” and “8” groups were significantly higher compared with that of the “0–2” group (all, p<0.001). The p for trend of the four groups was also significant (p<0.001) (Tables 2, 3).

**Table 2.** UWES-9 score of each group according to the number of workplace IPC measures for COVID-19 at the follow-up.

| UWES-9 item | Group by # of workplace IPC measures |  |  |  |  |  |  |  |
| --- | --- | --- | --- | --- | --- | --- | --- | --- |
|  | “0–2” group<br>(n=2,510) |  | “3–5” group<br>(n=3,240) |  | “6–7” group<br>(n=3,295) |  | “8” group<br>(n=2,937) |  |
|  | Mean | (SD) | Mean | (SD) | Mean | (SD) | Mean | (SD) |
| Total score | 18.3 | (13.2) | 20.8 | (12.8) | 21.7 | (12.2) | 22.6 | (12.6) |
| Vigor | 5.7 | (4.5) | 6.4 | (4.4) | 6.8 | (4.3) | 7.1 | (4.4) |
| Dedication | 6.7 | (4.7) | 7.7 | (4.5) | 8.0 | (4.3) | 8.2 | (4.4) |
| Absorption | 5.9 | (4.5) | 6.7 | (4.5) | 6.9 | (4.2) | 7.2 | (4.3) |

IPC, infection prevention and control; SD, standard deviation; UWES-9, nine-item version of the Utrecht Work Engagement Scale.

**Table 3.** Association between participants’ work engagement and number of workplace IPC measures for COVID-19.

| UWES-9 item<br>Group by # of<br>workplace IPC measures | Sex-age adjusted |  |  | Multivariate |  |  |
| --- | --- | --- | --- | --- | --- | --- |
|  | Coef. | [95%CI] | p | Coef. | [95%CI] | p |
| Total score |  |  |  |  |  |  |
| “0–2” group | Ref. |  | <0.001† | Ref. |  | <0.001† |
| “3–5” group | 2.59 | [1.94 – 3.24] | <0.001 | 2.86 | [2.20 – 3.52] | <0.001 |
| “6–7” group | 3.47 | [2.82 – 4.12] | <0.001 | 4.01 | [3.32 – 4.69] | <0.001 |
| “8” group | 4.34 | [3.67 – 5.01] | <0.001 | 5.04 | [4.32 – 5.77] | <0.001 |
| Vigor |  |  |  |  |  |  |
| “0–2” group | Ref. |  | <0.001† | Ref. |  | <0.001† |
| “3–5” group | 0.76 | [0.53 – 0.98] | <0.001 | 0.85 | [0.62 – 1.08] | <0.001 |
| “6–7” group | 1.06 | [0.84 – 1.29] | <0.001 | 1.25 | [1.01 – 1.49] | <0.001 |
| “8” group | 1.43 | [1.20 – 1.66] | <0.001 | 1.67 | [1.42 – 1.92] | <0.001 |
| Dedication |  |  |  |  |  |  |
| “0–2” group | Ref. |  | <0.001† | Ref. |  | <0.001† |
| “3–5” group | 1.02 | [0.79 – 1.25] | <0.001 | 1.11 | [0.88 – 1.34] | <0.001 |
| “6–7” group | 1.39 | [1.16 – 1.62] | <0.001 | 1.57 | [1.33 – 1.81] | <0.001 |
| “8” group | 1.57 | [1.34 – 1.81] | <0.001 | 1.82 | [1.56 – 2.07] | <0.001 |
| Absorption |  |  |  |  |  |  |
| “0–2” group | Ref. |  | <0.001† | Ref. |  | <0.001† |
| “3–5” group | 0.81 | [0.58 – 1.03] | <0.001 | 0.90 | [0.67 – 1.13] | <0.001 |
| “6–7” group | 1.02 | [0.79 – 1.24] | <0.001 | 1.19 | [0.95 – 1.42] | <0.001 |
| “8” group | 1.34 | [1.11 – 1.57] | <0.001 | 1.55 | [1.30 – 1.80] | <0.001 |

CI, confidence interval; IPC, infection prevention and control; UWES-9; nine-item version of the Utrecht Work Engagement Scale. † p for trend.

As for the mean (SD) subscale scores of vigor, dedication, and absorption among the four groups, the “0–2” group was again the lowest at 5.7 (4.5), 6.7 (4.7), and 5.9 (4.5), respectively. The “8” group was highest the highest at 7.1 (4.4), 8.2 (4.4), and 7.2 (4.3), respectively. In both the sex-age adjusted model and the multivariate model, the score of the of “3–5,” “6–7,” and “8” groups were significantly higher compared with that of the “0–2” group (all, p<0.001). The p for trend of the four groups was also significant (p<0.001) (Tables 2, 3).

## Discussion

In this study, using the data from the CORoNaWork Project, we analyzed how the number of workplace IPC measures implemented in the workplace affected the participants’ work engagement at their one-year follow-up. The results showed that there was an association between the implementation of workplace IPC measures and work engagement. In addition, we found that the more the workplace IPC measures were implemented, the higher the employees’ work engagement.

Since the COVID-19 pandemic in Japan began in March 2020, the policy of workplaces regarding IPC measures was considered to have been established and relayed to the employees at the time of the baseline survey (December 2020). During the one-year period between the baseline and the follow-up survey, there were three waves of the COVID-19 pandemic, during which we believe these IPC measures could have been implemented on a sustained basis. In addition, work engagement has been shown to be a sustained and general feeling, rather than a temporary and transient feeling towards work.^15, 16, 22^ Thus, we speculate that workplace IPC measures could have a sustained and generally positive effect on workers’ mental health.

There are several reasons why proactive workplace IPC measures may result in high work engagement among workers. Certain studies have reported that anxiety and fear of COVID-19 infection have directly led to negative mental health.^24-26^ In addition, we have reported that the more the workplace IPC measures are implemented in a workplace, the lower the psychological distress among workers.^27^ Workplace IPC measures may contribute to improved work engagement by decreasing employees’ anxiety and mental stress. However, it is unclear whether these IPC measures are effective in reducing the risk of actual COVID-19 infection; another study is needed to clarify this research question.

Workplace IPC measures can mainly be promoted using a top-down process, that is, through a management system wherein actions are initiated at the highest level. It has been suggested that a strong top-down process promotes a safe climate as well as workers’ psychosocial safety in the workplace, and contributes to the reduction of mental distress among workers.^28^ A previous study has reported that during the COVID-19 pandemic, the higher workers’ perceived workplace health support—that is, the support for workers’ lively working and healthy living provided by the workplace—the higher the health-related quality of life.^29^ In addition, clear policies surrounding workplace IPC measures have been reported to build trust between employers and workers.^30^ Actively promoting workplace IPC measures could also be found to enhance corporate governance, increase employees’ perceived workplace health support, and contribute to positive mental health, including increased work engagement.

Workplace IPC measures tend to be implemented more in the secondary industry than in the tertiary industry. Those who work for companies that implement more workplace IPC measures have been reported to be more well-educated and belong to large-sized companies.^27, 31^ These may result in various occupational factors, such as the difficulty in introducing workplace IPC measures in certain industries; the influence of risk awareness among management and employers; the presence or absence of interventions by occupational health specialists, such as occupational physicians and occupational hygienists; and the costs associated with implementing the countermeasures.

## Limitations

This study has several limitations. First, the present study only included as participants those who were registered as Internet-based survey monitors. Therefore, the sample of the study may not represent the general working population, and the generalizability of this study should be treated with caution. For example, there is a risk of overestimation if multiple participants belong to the same workplace. To deal with such issues, we made an effort to reduce sample bias by conducting random sampling stratified by gender and region of residence. Second, we did not consider the continuity or intensity of implementation of the workplace IPC measures. The baseline survey for this study was conducted in December 2020, when the third wave of the COVID-19 pandemic was expanding nationwide. Therefore, we assumed that workplace IPC measures were implemented with high intensity. However, the intensity of the participants’ self-IPC measures and employees’ perceptions thereof could differ between the period where the pandemic was under control and the period where the third wave was expanding. Third, workplaces that implement many IPC measures may have ordinarily been engaged in health and productivity management, implementing workers’ mental health measures, or concerned about the well-being of their employees. Thus, the participants may have already had less mental distress at baseline. However, it is difficult to evaluate this point in the present study.

## Conclusions

This study found that promoting workplace IPC measures improved workers’ work engagement. It was also shown that a dose-response relationship existed between workplace IPC measures and work engagement. Workplace IPC measures are expected to reduce workers’ fear and anxiety related to COVID-19 infection and to contribute to the mental health of workers. We believe that the implementation of workplace IPC measures could be important not only in controlling the COVID-19 pandemic, but also in promoting the positive mental health of workers.

## Data Availability

All data produced in the study are available upon reasonable request to the authors.

## Author contributions

K.I. wrote the manuscript and analyzed the data; H.A. analyzed the data; and Y.F. was the chairperson of the study group. All the authors designed the research protocol and developed the questionnaire. All authors reviewed and approved the final manuscript.

## Acknowledgements

This study was supported and partly funded by the research grant from the University of Occupational and Environmental Health, Japan (no grant number); Japanese Ministry of Health, Labour and Welfare (H30-josei-ippan-002, H30-roudou-ippan-007, 19JA1004, 20JA1006, 210301-1, and 20HB1004); Anshin Zaidan (no grant number), the Collabo-Health Study Group (no grant number), and Hitachi Systems, Ltd. (no grant number) and scholarship donations from Chugai Pharmaceutical Co., Ltd. (no grant number). The funders were not involved in the study design, collection, analysis, interpretation of data, the writing of this article or the decision to submit it for publication. The current members of the CORoNaWork Project, in alphabetical order, are as follows: Yoshihisa Fujino (present chairperson of the study group), Akira Ogami, Arisa Harada, Ayako Hino, Hajime Ando, Hisashi Eguchi, Kazunori Ikegami, Kei Tokutsu, Keiji Muramatsu, Koji Mori, Kosuke Mafune, Kyoko Kitagawa, Makoto Okawara, Masako Nagata, Mayumi Tsuji, Ning Liu, Rie Tanaka, Ryutaro Matsugaki, Seiichiro Tateishi, Shinya Matsuda, Tomohiro Ishimaru, and Tomohisa Nagata. All members are affiliated with the University of Occupational and Environmental Health, Japan. Finally, we would like to thank Editage (http://www.editage.com) for English-language editing.

## Data availability statement

All data produced in the study are available upon reasonable request to the authors.

## Disclosure

### Ethical approval

This study was approved by the ethics committee of the University of Occupational and Environmental Health, Japan (Reference No. R2-079 and R3-006).

### Informed consent

Informed consent from all participants was obtained via website.

### Registry and registration no. of the study/trial

N/A.

### Animal studies

N/A.

### Conflict of interest

Authors declare no Conflict of Interests for this article.

## Notes

### Competing Interest Statement

The authors have declared no competing interest.

### Author Declarations

The ethics committee of the University of Occupational and Environmental Health, Japan gave ethical approval for this work(Reference No. R2-079 and R3-006).

